# Country-level Determinants of the Severity of the First Global Wave of the COVID-19 Pandemic: An Ecological Study

**DOI:** 10.1101/2020.05.13.20100677

**Authors:** Tiberiu A Pana, Sohinee Bhattacharya, David T Gamble, Zahra Pasdar, Weronika A Szlachetka, Jesus A Perdomo-Lampignano, Kai D Ewers, David J McLernon, Phyo K Myint

**Affiliations:** Institute of Applied Health Sciences, School of Medicine, Medical Sciences & Nutrition, University of Aberdeen, United Kingdom

## Abstract

**Objective:** We aimed to identify the country-level determinants of the severity of the first wave of the COVID-19 pandemic.

**Design:** An ecological study design of publicly available data was employed. Countries reporting >25 COVID-related deaths until 08/06/2020 were included. The outcome was log mean mortality rate from COVID-19, an estimate of the country-level daily increase in reported deaths during the ascending phase of the epidemic curve. Potential determinants assessed were most recently published demographic parameters (population and population density, percentage population living in urban areas, median age, average body mass index, smoking prevalence), Economic parameters (Gross Domestic Product per capita); environmental parameters: pollution levels, mean temperature (January-May)), co-morbidities (prevalence of diabetes, hypertension and cancer), health system parameters (WHO Health Index and hospital beds per 10,000 population); international arrivals, the stringency index, as a measure of country-level response to COVID-19, BCG vaccination coverage, UV radiation exposure and testing capacity. Multivariable linear regression was used to analyse the data.

**Primary Outcome:** Country-level mean mortality rate: the mean slope of the COVID-19 mortality curve during its ascending phase.

**Participants:** Thirty-seven countries were included: Algeria, Argentina, Austria, Belgium, Brazil, Canada, Chile, Colombia, the Dominican Republic, Ecuador, Egypt, Finland, France, Germany, Hungary, India, Indonesia, Ireland, Italy, Japan, Mexico, the Netherlands, Peru, the Philippines, Poland, Portugal, Romania, the Russian Federation, Saudi Arabia, South Africa, Spain, Sweden, Switzerland, Turkey, Ukraine, the United Kingdom and the United States.

**Results:** Of all country-level predictors included in the multivariable model, total number of international arrivals (beta 0.033 (95% Confidence Interval 0.012,0.054)) and BCG vaccination coverage (−0.018 (−0.034,-0.002)), were significantly associated with the mean death rate.

**Conclusions:** International travel was directly associated with the mortality slope and thus potentially the spread of COVID-19. Very early restrictions on international travel should be considered to control COVID outbreak and prevent related deaths.

**ARTICLE SUMMARY:** *Strengths and limitations:*

- A comparable and relevant outcome variable quantifying country-level increases in the COVID-19 death rate was derived which is largely independent of different testing policies adopted by each country
- Our multivariable regression models accounted for public health and economic measures which were adopted by each country in response to the COVID-19 pandemic by adjusting for the Stringency Index
- The main limitation of the study stems from the ecological study design which does not allow for conclusions to be drawn for individual COVID-19 patients
- Only countries that had reported at least 25 daily deaths over the analysed period were included, which reduced our sample and consequently the power.

## INTRODUCTION

The atypical pneumonia caused by SARS-CoV 2 has spread rapidly. As of the 8^th^ of June 2020, there have been over 400,857 deaths related to COVID-19 infection worldwide.^1^ The estimated overall case fatality rate is ∼7%, with country-level estimates ranging between 0.5-14%.^2^ Nevertheless, there is wide variation in the reported country-specific death rates which may be attributed to variation in testing rates, underreporting or real differences in environmental, sociodemographic and health system parameters.

Country-level determinants of the pandemic severity are largely unknown. The only previous ecological study to date assessing country-level predictors of the severity of the COVID-19 pandemic including data on 65 countries^3^ has found that the cumulative number of infected patients in each country was directly associated with the case fatality rate, whilst testing intensity was inversely associated with case fatality rate. This study found no association between health expenditure and case fatality rate. However, other important country-level determinants were not evaluated and thus their relationship with pandemic severity remains unknown.

Several risk factors for COVID-related mortality have been proposed, including older population,^4^ higher population co-morbid burden,^5^ smoking,^6^ obesity,^7^ pollution levels^8^ and healthcare system performance.^9^ Furthermore, countries outside China most severely hit by the pandemic were those with a high income, high GDP per capita and well-established healthcare systems, such as Italy, Spain, France, the United Kingdom and the United States.^10^ In contrast, lower- and middle-income countries reported much lower COVID-19 incidence and mortality rates.^10^ Whilst these differences may be attributable to case under-reporting and infrequent testing in these countries, other factors may also be involved.

In this study, we aimed to assess the country-level determinants of the severity of the first wave of the COVID-19 pandemic based on currently available evidence using publicly available data and an ecological study design.

## METHODS

### Patient and Public Involvement

There was no patient or public involvement in designing the study given the urgent nature of the COVID-19 pandemic and the usage of publicly available data.

### Study Design

An ecological study design was used. The outcome was the steepness of the ascending curve of country specific daily reports of COVID-19 related deaths between 31/12/2019-08/06/2020. The following determinants were assessed: demographic predictors (population and population density, percentage population living in urban areas, proportion of population aged 65 and over, average body mass index (BMI), smoking prevalence), economic predictors (gross Domestic Product (GDP) per capita), environmental predictors (pollution levels, mean temperature (January-May) [2010-2016]), prevalent co-morbidities (diabetes, hypertension and cancer), health systems predictors (WHO Health Index and hospital beds per 10,000 population), international arrivals (as a proxy measure of the globalisation status of each country), the stringency index (as measure of country level response to the pandemic)^11^, exposure to UV radiation (as a proxy for sunlight exposure), BCG vaccination coverage and testing capacity.

### Ethics Committee Approval

Given the study design and the use of publicly available data, no ethical approval was considered necessary.

### Selection criteria

Countries reporting at least 25 daily deaths up to the 8^th^ of June 2020 with available data for all chosen determinants were included. A total of 37 countries from 4 continents were included in the analysis: Africa (Algeria, Egypt, South Africa), America (Argentina, Brazil, Canada, Chile, Colombia, the Dominican Republic, Ecuador, Mexico, Peru and the United States of America), Asia (India, Indonesia, Japan, the Philippines, Saudi Arabia, Turkey) and Europe (Austria, Belgium, Finland, France, Germany, Hungary, Ireland, Italy, the Netherlands, Poland, Portugal, Romania, the Russian Federation, Spain, Sweden, Switzerland, Ukraine, the United Kingdom). China was not included in the analysis due to potential inaccuracies in the number of daily reported deaths which may have occurred subsequent to 1290 deaths which were retrospectively reported on the 17^th^ of April.^12^

### Data Sources

Country-level parameters were obtained from freely accessible data sources. The daily reported number of COVID-19 cases and deaths between 31/12/2019-08/06/2020 as well as the 2018 population data were extracted from the European Centre for Disease Control.^13^

The data regarding the median population age and population density were extracted from the United Nations World Population Prospects^14^ and United Nations Statistics Division, respectively.^15^ The data regarding the percentage of the population living in urban areas were extracted from the World Urbanisation Prospects, issued by the United Nations Population Division.^16^ Temperature data were extracted from the Climate Change Knowledge Portal from the World Bank Group.^17^ Prevalent diabetes, gross domestic product, international arrivals in 2018, and current health expenditure data were extracted from the World Development Indicators (WDI) database, provided by the World Bank Group.^18^ Data regarding prevalent cancers, proportion of population aged 65 and over and the total number of COVID-19 tests performed were extracted from the Our World in Data and the Sustainable Development Goals (SDG) tracker,^19, 20^ an open-access publication tracking global progress to the United Nations Sustainable Development Goals for global development, adopted in September 2015. Prevalent hypertension, body mass index (BMI), cigarette smoking, ambient air pollution, ultraviolet (UV) radiation and Bacillus Calmette– Guérin (BCG) vaccination data were obtained from the Global Health Observatory (GHO) data repository of the World Health Organization.^21^ The world health organisation health index was extracted from the WHO Global Partnership for Education (GPE) paper series published in 2000.^22^ Country-level total hospital beds per 10,000 population data were extracted from the World Bank Dataset “World Bank Indicators of Interest to the COVID-19 Outbreak”.^23^ Daily Stringency Index (SI) measurements between 31/01/2019-08/06/2020 were extracted from the Oxford COVID-19 Government Response Tacker (OxCGRT).^11^

### Definition of outcome and predictors

#### Outcome

Whilst previous ecological studies of other epidemics have utilised case or death counts as outcome,^24^ this may be prone to bias due to variations in country-level testing strategies,^25^ variations in population movement controls and differences in secondary attack rates within community cohorts^26^. The mean mortality rate was thus chosen as outcome instead, since it is independent of these parameters and may thus represent a more reliable indicator of the country-level severity of the COVID-19 pandemic

Mean mortality rate was defined as the mean slope of the mortality curve (Figure 1), measured from the first day when more than 2 COVID-19 deaths were reported until either the mortality curve reached a peak value or the 8^th^ of June 2020, whichever occurred first. The peak of each mortality curve was defined as the first point at which the first derivate of the COVID-19 mortality as a function of the pandemic timeline became zero. Before slope calculation, the mortality curve in each country was smoothed using a locally weighted (Lowess) regression using a bandwidth of 0.4. In order to ensure a good fit of the Lowess regression line, only countries having reported at least 25 daily deaths until the 8^th^ of June 2020 were included. The mean mortality rate thus represents an estimate of the country-level daily increase in reported deaths during the ascending phase of the epidemic curve.

**Figure 1.**
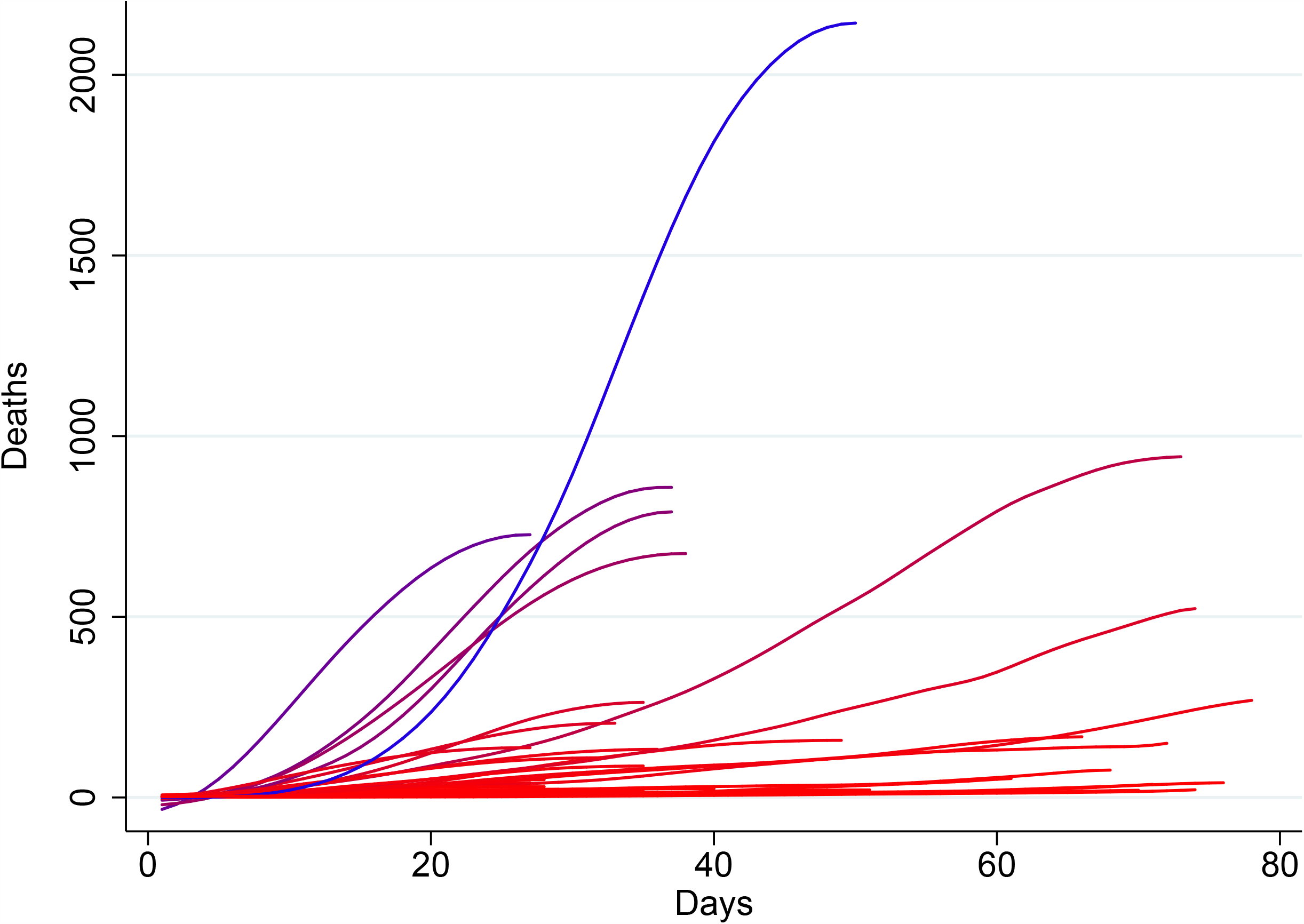
Graphical representation of the smoothed* number of daily deaths of each country (before reaching mortality peak, if applicable) as a function of the number of days passed since the first day when an excess of 3 deaths were reported. Countries with higher mortality rates are depicted in blue, while those with lower mortality rates are depicted in red. *smoothed using a local regression (lowess) function with a bandwidth of 0.4

#### Determinants

Data on population density were extracted as the country-level population per square kilometre in 2019.^27^ Data on ambient air pollution were extracted as the country-level mean concentration of fine particulate matter (PM2.5) measured in 2016.^28^ Temperature data were extracted as the mean temperature recorded in each country between January and May using temperature data recorded between 2010 and 2016.^17^ Data on International Arrivals were extracted as the total number of country-level international arrivals in 2018.^29^

Data on prevalent diabetes were extracted as the percentage of the population aged 20 to 79 years in 2019.^18^ Data on prevalent cancers were extracted as the age-standardized cancer prevalence among both sexes in 2017, expressed as percentages.^30^ Data on prevalent hypertension were extracted as the age-standardised percentage of the population over 18 years of age with systolic blood pressure ≥140 mmHg or diastolic blood pressure ≥90 mmHg in 2015.^31^ Data on BMI were extracted as the age-standardised mean body mass index trend estimates for both sexes amongst adults (≥18 years) in 2016.^32^ Data on daily cigarette smoking were extracted as the age-standardised smoking rate across both sexes amongst adults (≥18 years) in 2013.^33^ Whilst the definition of “daily cigarette smoking” varies across surveys, it habitually refers to current smoking of cigarettes at least once a day.^33^

Data on GDP were extracted as GDP per capita by Purchasing Power Parity (PPP) in current international dollars in 2018.^34^ The percentage of population living in urban areas was defined as the percentage of de facto population living in areas classified as urban according to the criteria used by each area or country.^16^ The World Health Organisation (WHO) health index is a composite index that aims to evaluate a given countries healthcare system performance relative to the maximum it could achieve given its level of resources and non-healthcare system determinants. It was calculated in the year 2000. The index uses five weighted parameters: overall or average disability-adjusted life expectancy (25%), distribution or equality of disability-adjusted life expectancy (25%), overall or average healthcare system responsiveness (including speed of provision and quality of amenities; 12.5%), distribution or equality of healthcare system responsiveness (12.5%) and healthcare expenditure (25%). Data on hospital beds per 10,000 population were defined by the World Bank as including ‘inpatient beds available in public, private, general, and specialized hospitals and rehabilitation centres’. The published data for countries included was from 2000 to 2017. In most cases beds for both acute and chronic care are included.^23^ The Stringency Index is an overall indicator of public health measures adopted by each country in response to the COVID-19 pandemic and includes containment and closure indicators (school closures, workplace closures, cancelling public events, restrictions on gatherings, public transport closures, stay-at-home requirements, restrictions on internal movements, international travel controls), economic response indicators (income support, debt/contract relief, fiscal measures, international support) as well as health systems indicators (public information campaigns, testing policy, contact tracing, emergency investment in healthcare, investment in vaccines).^11^ The mean daily Stringency Index was calculated for each country between 31/12/2019 and until either the mortality curve reached a peak value or the 8^th^ of June 2020, whichever occurred first.

Country-level exposure to UV radiation was quantified as the population-weighted average daily ambient ultraviolet radiation level measured in J/m^2^ for the years 1997-2003.^35^ BCG vaccination coverage was quantified as the average percentage of 1 year-old children having received the BCG vaccine between 1980 and 2019 in each country. Testing capacity was quantified as the total number of COVID-19 tests per 1000 population performed until the 8^th^ of June 2020.

### Statistical analysis

All analyses were performed in Stata 15.1SE, Stata Statistical Software. A 5% threshold of statistical significance was utilised for all analyses (*P* <0.05). Linear regressions were performed to assess the univariable relationship between each country-level predictor and the calculated mean mortality rate for each country. The following predictors were included in the univariable analyses: the natural logarithm of the population in 2018 (10 million incraese), percentage of population aged 65 and over, pollution levels, mean temperature (January-May), international arrivals, population density, prevalent diabetes, prevalent neoplasms, median BMI, prevalent hypertension, smoking prevalence, hospital beds (per 10,000 population), WHO health index, percentage population living in urban areas, GDP per capita (PPP), UV radiation exposure, mean BCG coverage and the stringency index. Predictors reaching a *P*-value <0.3 at univariable level were then included in a multivariable logistic regression model with the natural logarithm of the mean mortality rate as outcome: the logarithm of the total population in 2018, percentage of population aged 65 and over, pollution, mean temperature (January-May), international arrivals, population density, prevalent neoplasms, prevalent hypertension, the WHO health index, population living in urban areas, GDP per capita, UV radiation exposure, mean BCG coverage and the stringency index.

Given that testing capacity data for 8 (Algeria, Brazil, Egypt, France, Germany, the Netherlands, Spain and Sweden) of the 37 included countries were not available, a secondary analysis also including testing capacity as a predictor was performed considering only the remaining 29 countries. Linear regressions were performed to assess the univariable relationship between each country-level predictor and the calculated mean mortality rate for each country. Predictors reaching a *P*-value <0.3 at univariable level were then included in a multivariable logistic regression model with the natural logarithm of the mean mortality rate as outcome: the logarithm of the total population in 2018, percentage of population aged 65 and over, international arrivals, population density, prevalent neoplasms, prevalent hypertension, GDP per capita, UV radiation exposure, mean BCG coverage, the stringency index and testing capacity.

## RESULTS

Table 1 and Supplementary File 1 detail the analysed data for the 37 included countries, including the calculated mean mortality rates. The mean mortality rates ranged between 0.22 (Chile) and 43.74 (the United States) new daily deaths. Only five included countries had a high mean mortality rate (>10): the United States (43.74), Spain (29.23), the United Kingdom (24.05), France (22.13), Italy (18.79) and Brazil (13.09).

**Table 1.** Observed mean mortality rate and number of international arrivals in 2018 (millions) for each country included in the analyses. Countries were categorised in 3 groups: high mean mortality rate group (>20 additional daily deaths), medium mean mortality rate group (2-20 additional daily deaths) and low mean mortality rate group (<2 additional daily deaths).

| Country Name | Mean Mortality Rate<br>(daily increase in deaths)<br>[up to 01/05/20] | International Arrivals<br>(millions) [2018] |
| --- | --- | --- |
| **High Mean Mortality Rate** |  |  |
| United States of America | 43.74 | 79.75 |
| Spain | 29.23 | 82.77 |
| United Kingdom | 24.05 | 36.32 |
| France | 22.13 | 89.32 |
| Italy | 18.79 | 61.57 |
| Brazil | 13.09 | 6.62 |
| **Medium Mean Mortality Rate** |  |  |
| Belgium | 7.86 | 9.12 |
| Mexico | 7.15 | 41.31 |
| Germany | 6.58 | 38.88 |
| Netherlands | 5.40 | 18.78 |
| Turkey | 3.48 | 45.77 |
| India | 3.48 | 17.42 |
| Canada | 3.27 | 21.13 |
| Sweden | 2.59 | 7.44 |
| Russian Federation | 2.52 | 24.55 |
| Peru | 2.05 | 4.42 |
| **Low Mean Mortality Rate** |  |  |
| Switzerland | 1.60 | 10.36 |
| Ireland | 1.58 | 10.93 |
| Portugal | 1.03 | 16.19 |
| Algeria | 0.88 | 2.66 |
| South Africa | 0.84 | 10.47 |
| Ecuador | 0.81 | 2.54 |
| Poland | 0.79 | 19.62 |
| Indonesia | 0.72 | 15.81 |
| Austria | 0.70 | 30.82 |
| Romania | 0.60 | 11.72 |
| Egypt | 0.50 | 11.20 |
| Japan | 0.48 | 31.19 |
| Saudi Arabia | 0.48 | 15.33 |
| Philippines | 0.46 | 7.17 |
| Colombia | 0.42 | 3.90 |
| Hungary | 0.38 | 17.55 |
| Ukraine | 0.31 | 14.10 |
| Dominican Republic | 0.28 | 6.57 |
| Finland | 0.26 | 3.22 |
| Argentina | 0.25 | 6.94 |
| Chile | 0.22 | 5.72 |

Table 2 details the results of the linear regression analyses. The following country-level predictors showed a statistically significant relationship with log mean mortality rate at univariable level: natural logarithm of population, international arrivals, prevalent neoplasms, prevalent hypertension, GDP per capita and BCG vaccination coverage. Upon multivariable adjustment, International arrivals in 2018, as a marker of global connection, was the main statistically significant predictor of log mean mortality rate (0.040 (0.017, 0.063) for 1 million increase in international arrivals, *P* =0.002) along with mean BCG vaccination coverage (−0.018 (−0.034, -0.002) for 1% increase in BCG vaccination coverage, *P* =0.031). Figures 2 and 3 detail the relationship between the country-level log mean mortality rate (predicted and observed) and each country-level predictor included in the multivariable regression model.

**Table 2.** Results of the linear regression assessing the country-level predictors of the daily increase in deaths. The predictors achieving a 30% statistical significance level at univariable levels (*P* < 0.3) were included in the multivariable model.

| Predictor | Univariable |  | Multivariable |  |
| --- | --- | --- | --- | --- |
|  | Coefficient (95% CI) | *P* value | Coefficient (95% CI) | *P* value |
| Natural logarithm of population (10 million increase) [2018] | **0.432 (0.050, 0.814)** | **0.033** | 0.393 (-0.087, 0.873) | 0.103 |
| % population aged 65 and older | 0.065 (-0.010, 0.139) | 0.097 | -0.020 (-0.143, 0.103) | 0.741 |
| Pollution levels | -0.017 (-0.044, 0.011) | 0.247 | -0.005 (-0.031, 0.020) | 0.659 |
| Mean Temperature (January-May) [2010-2016] | -0.031 (-0.078, 0.017) | 0.218 | 0.052 (-0.025, 0.128) | 0.175 |
| International Arrivals (1 million increase) [2018] | **0.049 (0.033, 0.064)** | **<0.001** | **0.033 (0.012, 0.054)** | **0.003** |
| Population Density | -0.002 (-0.006, 0.002) | 0.268 | -0.001 (-0.004, 0.002) | 0.560 |
| Diabetes prevalence (% of population ages 20 to 79) [2019] | -0.0031 (-0.189, 0.126) | 0.700 | - | - |
| Prevalence - Neoplasms - Sex: Both - Age: Age-standardized (Percent) (%) [2017] | **0.614 (0.209, 1.019)** | **0.005** | -0.404 (-1.079, 0.271) | 0.227 |
| Median BMI | 0.010 (-0.297, 0.318) | 0.947 | - | - |
| Prevalent Hypertension (%), [2015] | **-0.150 (-0.254, -0.045)** | **0.008** | -0.107 (-0.249, 0.035) | 0.132 |
| Smoking prevalence, 2016 total (ages 15+) | 0.002 (-0.058, 0.062) | 0.952 | - | - |
| Hospital beds (per 10, 000 population) | -0.004 (-0.022, 0.014) | 0.632 | - | - |
| WHO health index, [2000] | 2.259 (-0.920, 5.439) | 0.173 | -2.616 (-6.157, 0.925) | 0.140 |
| Population living in urban areas (%) | 0.023 (-0.011, 0.580) | 0.193 | 0.010 (-0.019, 0.039) | 0.468 |
| GDP per capita, PPP (\$1000 increase), [2018] | **0.280 (0.037, 0.524)** | **0.030** | 0.154 (-0.174, 0.482) | 0.340 |
| Country-level average daily ambient ultraviolet radiation (UVR) level - 2004 | -0.000 (-0.001, 0.000) | 0.133 | -0.001 (-0.001, 0.000) | 0.109 |
| Mean % of BCG vaccination coverage among 1 year old children (1980-2019) | **-0.027 (-0.037, -0.016)** | **<0.001** | **-0.018 (-0.034, -0.002)** | **0.031** |
| Mean Daily Stringency Index | -0.036 (-0.072, 0.001) | 0.057 | 0.004 (-0.028, 0.037) | 0.790 |

$R^2$ for multivariable linear regression = 0.8031
BMI – body mass index; WHO – world health organisation; GDP – gross domestic product; PPP – purchasing power parity; BCG – Bacille-Calmette-Guerin

**Figure 2.**
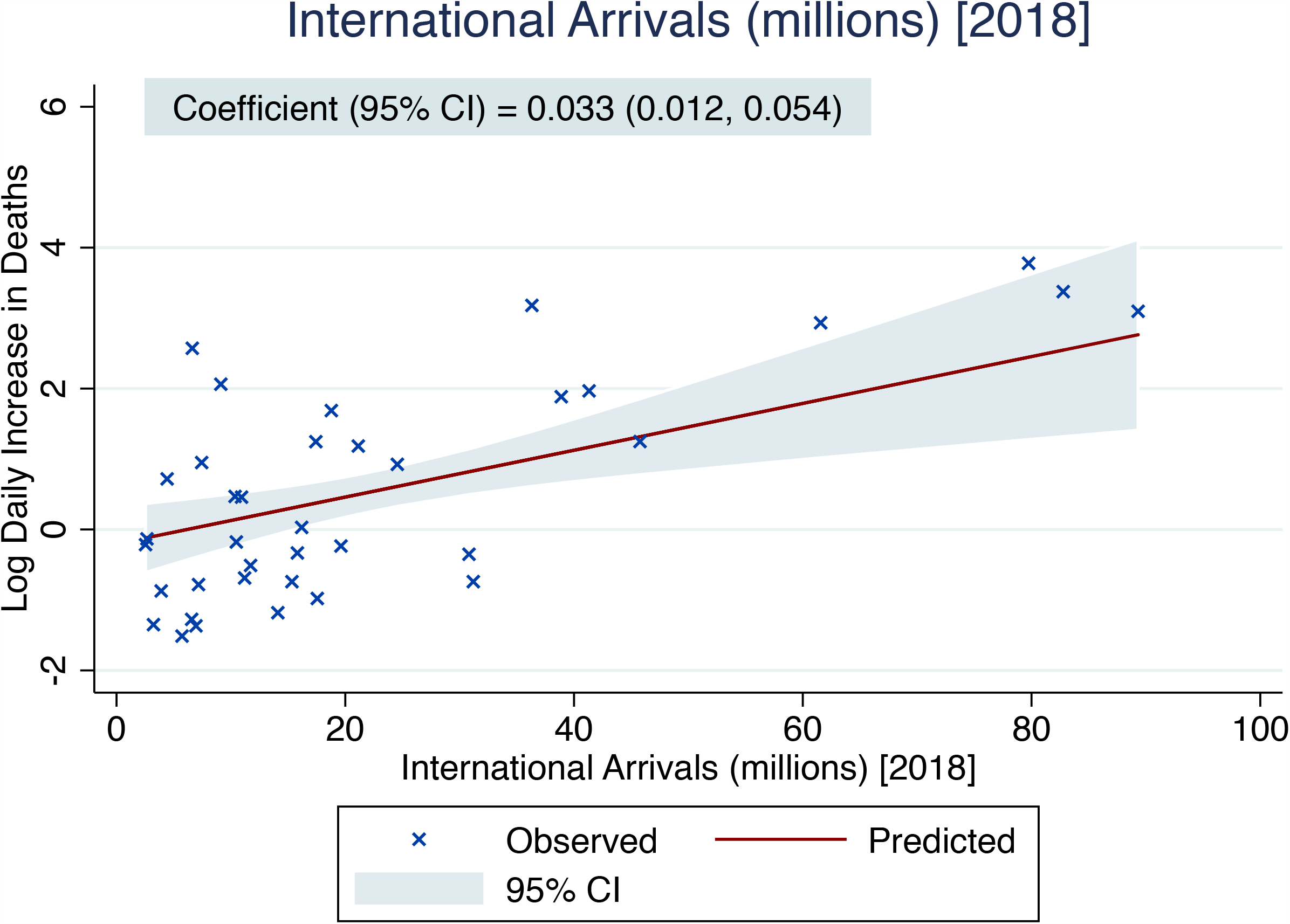
Predicted (based on the results of the multivariable linear regression) and observed country-level mortality rate (mean daily increase in deaths until the peak in mortality) as a function of the recorded country-level number of international arrivals in 2018 (millions). The solid red line represents the point estimate of the predicted log daily increase in deaths, while the blue-grey area represents the corresponding 95% confidence interval. The crosses represent the observed values of the log daily increase in deaths.

**Figure 3.**
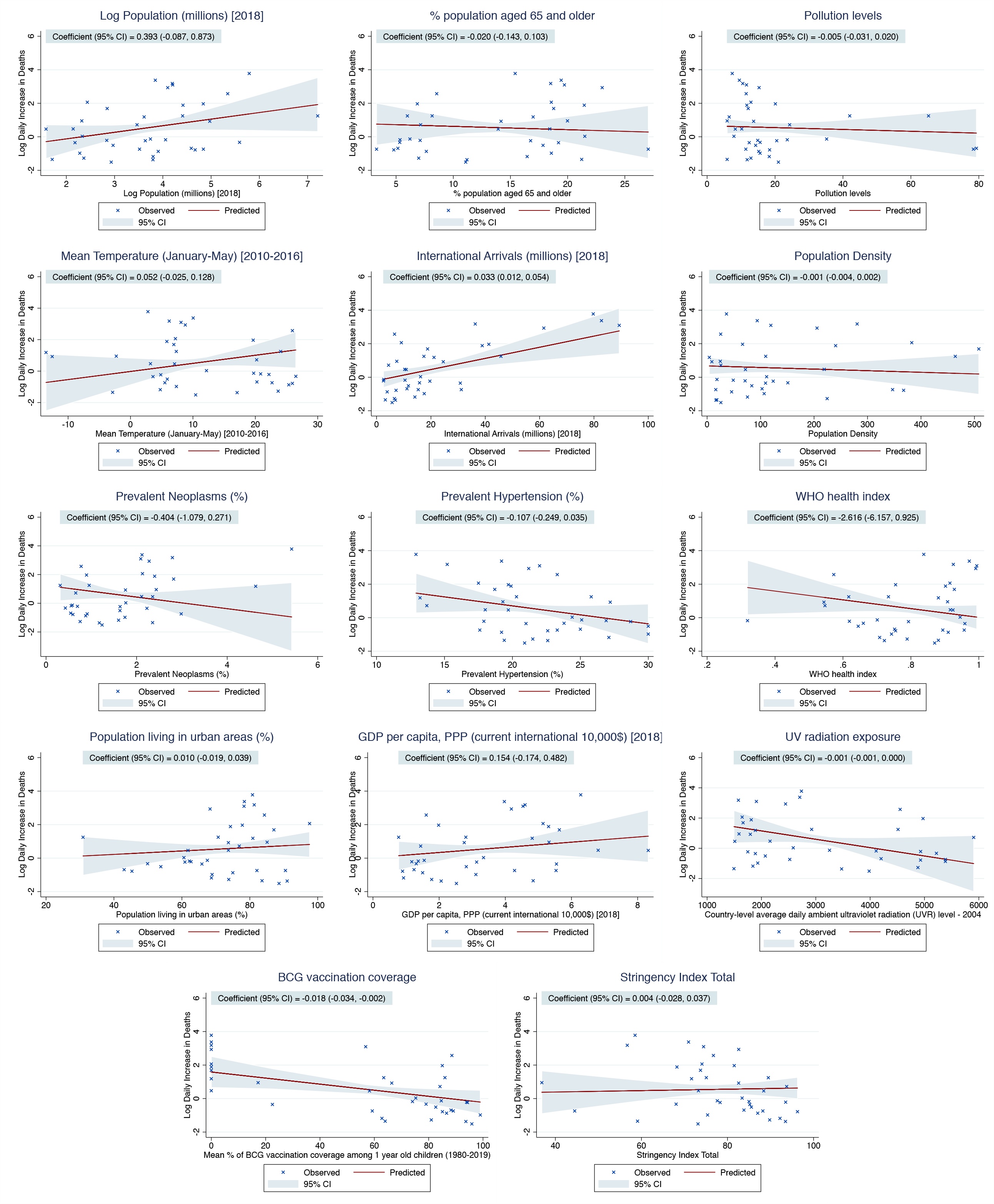
Predicted (based on the results of the multivariable linear regression) and observed country-level mortality rate (mean daily increase in deaths until the peak in mortality) as a function of each country-level predictor included in the multivariable model. The solid red lines represent the point estimates of the predicted log daily increase in deaths, while the blue-grey areas represent the corresponding 95% confidence intervals. The crosses represent the observed values of the log daily increase in deaths.

Table 3 details the results of the secondary linear regression analyses, including only countries having reported COVID-19 testing data up to the 8^th^ of June 2020. The following country-level predictors showed a statistically significant relationship with log mean mortality rate at univariable level: natural logarithm of population, international arrivals, prevalent neoplasms, prevalent hypertension, BCG vaccination coverage and total COVID-19 tests per 1000 population performed until the 8^th^ of June 2020. Upon multivariable adjustment, the statistically significant predictors of log mean mortality rate were: international arrivals in 2018 (0.036 (0.008, 0.063) for 1 million increase in international arrivals, *P* =0.013), prevalent hypertension (−0.129 (−0.246,-0.012) for 1% increase in country-level hypertension prevalence, *P* =0.032) and testing capacity (0.018 (0.001, 0.034) for 1 per 1000 population increase in the number of total COVID-19 tests performed until the 8^th^ of June 2020, *P* =0.039).

**Table 3.** Results of the secondary linear regression assessing the country-level predictors of the daily increase in deaths, including only countries reporting total COVID-19 tests performed up to the 8^th^ of June 2020. The predictors achieving a 30% statistical significance level at univariable levels (*P* < 0.3) were included in the multivariable model.

| Predictor | Univariable |  | Multivariable |  |
| --- | --- | --- | --- | --- |
|  | Coefficient (95% CI) | *P* value | Coefficient (95% CI) | *P* value |
| Natural logarithm of population (10 million increase) [2018] | **0.419 (0.038, 0.800)** | **0.040** | 0.385 (-0.044, 0.813) | 0.075 |
| % population aged 65 and older | 0.035 (-0.047, 0.118) | 0.407 | - | - |
| Pollution levels | -0.003 (-0.037, 0.030) | 0.848 | - | - |
| Mean Temperature (January-May) [2010-2016] | -0.032 (-0.081, 0.017) | 0.207 | 0.026 (-0.052, 0.104) | 0.484 |
| International Arrivals (1 million increase) [2018] | **0.059 (0.039, 0.079)** | **<0.001** | **0.036 (0.008, 0.063)** | **0.013** |
| Population Density | 0.002 (-0.002, 0.007) | 0.270 | 0.000 (-0.004, 0.003) | 0.822 |
| Diabetes prevalence (% of population ages 20 to 79) [2019] | 0.012 (-0.173, 0.196) | 0.903 | - | - |
| Prevalence - Neoplasms - Sex: Both - Age: Age-standardized (Percent) (%) [2017] | **0.582 (0.177, 0.987)** | **0.009** | -0.391 (-1.014, 0.233) | 0.203 |
| Median BMI | 0.107 (-0.205, 0.419) | 0.507 | - | - |
| Prevalent Hypertension (%), [2015] | **-0.140 (-0.240, -0.039)** | **0.011** | **-0.129 (-0.246, -0.012)** | **0.032** |
| Smoking prevalence, 2016 total (ages 15+) | -0.016 (-0.077, 0.045) | 0.610 | - | - |
| Hospital beds (per 10, 000 population) | -0.009 (-0.027, 0.009) | 0.323 | - | - |
| WHO health index, [2000] | 1.247 (-2.180, 4.675) | 0.482 | - | - |
| Population living in urban areas (%) | 0.007 (-0.030, 0.044) | 0.710 | - | - |
| GDP per capita, PPP (\$1000 increase), [2018] | 0.242 (-0.016, 0.499) | 0.077 | -0.045 (-0.325, 0.235) | 0.739 |
| Country-level average daily ambient ultraviolet radiation (UVR) level - 2004 | -0.000 (-0.001, 0.000) | 0.283 | 0.000 (-0.001, 0.000) | 0.310 |
| Mean % of BCG vaccination coverage among 1 year old children (1980-2019) | **-0.028 (-0.039, -0.017)** | **<0.001** | -0.011 (-0.029, 0.007) | 0.221 |
| Mean Daily Stringency Index | -0.033 (-0.074, 0.008) | 0.128 | 0.013 (-0.021, 0.048) | 0.425 |
| Total COVID-19 tests per 1000 population | **0.024 (0.008, 0.039)** | **0.007** | **0.018 (0.001, 0.034)** | **0.039** |
| (up to the 8<sup>th</sup> of June 2020) |  |  |  |  |

$R^2$ for multivariable linear regression = 0.8373
BMI – body mass index; WHO – world health organisation; GDP – gross domestic product; PPP – purchasing power parity; BCG – Bacille-Calmette-Guerin

## DISCUSSION

### Principal findings

In this ecological study including data from 37 countries which were most severely affected by COVID-19 in the first wave of current Global pandemic, we assessed 19 country-level socioeconomic, environmental, health and healthcare system, and globalisation parameters as potential predictors of variation in death rates from COVID 19 infection. In the multivariable linear regression model, the main predictor that reached statistical significance was international arrivals, a proxy of global connection: increases in international arrivals were associated with higher mean mortality rate. Furthermore, country-level BCG vaccination coverage was associated with decreases in the COVID-19 mean mortality rate during the first wave of the pandemic. Finally, in our secondary analyses including only country with available testing capacity data, the total number of COVID-19 tests performed per 1000 population until the 8^th^ of June 2020 was associated with increases in the COVID-19 mean mortality rate.

### Comparison with previous literature

A previous ecological study analysed the country-level predictors of the COVID-19 case fatality rate including 65 countries.^3^ This study found that upon adjustment for epidemic age, health expenditure and world region, the case fatality rate was significantly associated with increasing cumulative number of COVID-19 cases and decreasing testing intensity.^3^ Nevertheless, no other country-level predictors were included in this study.

Further comparisons can be made with data from previous pandemics. A negative association has been reported between health expenditure and death rates from the 2009 influenza pandemic in 30 European countries.^24^ Associations have also been reported between airline travel and spread of the H1N1 influenza virus infection.^36^

Comorbidities may account for mortality rate differences between countries. A study among laboratory-confirmed cases of COVID-19 in China showed that patients with any comorbidity, including diabetes, malignancy and hypertension, had poorer clinical outcomes than those without.^5^ We thus accounted for country-level data on a selection of key comorbidities which included prevalent diabetes mellitus, neoplasms, and hypertension. BMI ≥40kg/m2 has been identified as an independent risk factor for severe COVID-19 illness.^7^ Finally, a recent systematic review on 5 studies from China showed that smoking is likely associated with negative outcomes and progression of COVID-19.^6^

### Interpretation of findings

In our multivariate model, the main significant determinant of mortality was international arrivals. Travel restrictions and their effectiveness in containing respiratory virus pandemics remains a contentious subject. In 2007 the WHO published a protocol on ‘rapid operations to contain the initial emergence of pandemic influenza’, which included recommendations on travel restrictions.^37^ However, subsequent guidance advises such restrictions are not recommended once a virus has spread significantly.^38^ A recent systematic review of 23 studies that demonstrated limited impact of travel restrictions in the containment of influenza: internal travel restrictions delayed pandemic peak by approximately 1.5 weeks, while 90% air travel restriction delayed the spread of pandemics by approximately 3–4 weeks but only reduced attack rates by less than 0.02%.^39^ However, another systematic review of combination strategies for pandemic influenza response showed that combination strategies including travel restrictions increased the effectiveness of individual policies.^40^

The WHO recommendations for pandemic preparedness and resilience suggest that points of entry into the country should be monitored by focussing on surveillance and risk communication to travellers but falls short of closing down international travel.^41^

Interestingly, during the COVID-19 pandemic, some countries such as Thailand have adopted aggressive international travel screening and isolation policies, which may have led to lower infection rates.^42^ Our study suggests that travel restrictions have the potential to influence the impact of the COVID-19 pandemic and should be considered as part of a structured and rapidly instigated pandemic preparedness plan. Furthermore, the mean stringency index, which also accounts for international travel restrictions amongst other measures, was not associated with the mean mortality rate in the multivariable model. This suggest that international travel restrictions and other containment measures may have been imposed too late to influence the steepness of the mortality curve and that the level of global connectivity of each country may influence the course of the epidemic mortality curve before the number of COVID-19 related cases and deaths reaches worrying levels.

Our multivariable model also suggests an inverse relationship between BCG vaccination coverage the mean mortality rate, in which increasing BCG vaccination coverage was associated with decreased mean mortality rate. The relationship between BCG vaccination and the evolution of the COVID-19 transmission and disease severity remains controversial.^43, 44^ While the BCG vaccine has been postulated to exhibit non-specific immunomodulatory properties, which may reduce SARS-CoV-2 viraemia after exposure,^43^ current epidemiological evidence is derived from ecological studies^45^ and needs to be interpreted in the light of the inherent limitations of this study design. Further ongoing studies (NCT04327206^46^, NCT04328441^47^) may provide more robust evidence regarding the association between BCG vaccination and COVID-19.

Our analyses also revealed a few surprising findings: the intensity of COVID-19 testing was apparently associated with mean mortality rate increases while the country-level prevalence of hypertension was apparently associated with mean mortality rate decreases. These findings appear to be contradictory to previous evidence suggesting that testing intensity may be associated with decreased COVID-19 mortality,^48^ while hypertension was clearly associated with increased mortality.^49^ These surprising findings need to be interpreted in the light of our ecological study design in which residual confounders may influence these associations.

### Strengths and Limitations

The main strength of this study lies in its use of comparable and relevant outcome data derived from contemporary death reporting from countries affected by COVID-19. As testing rates for the virus vary across countries, the incidence or prevalence of the disease cannot be compared between countries. While death from the disease is a hard outcome, the denominator information to calculate death rates make between-country comparisons difficult. In addition, the deaths in the community, particularly in the elderly living in care homes, often go untested and thus firm diagnosis remains impossible. Therefore, in this study we have adopted an outcome that is comparable in terms of the increase in the rate of death, rather than death rates *per se*. Therefore, this may better represent the spread and seriousness of pandemic in individual countries when comparing countries at different stages of the pandemic. The country-level parameters assessed as potential predictors have all been implicated at some point to be associated with severity and consequently mortality. We however found that the main predictor of the total number of international arrivals in the country (2018 figures), signifying transmission of the infection through travel. Although the data was from 2018, there is no reason to believe that international travel figures between countries would be different in early 2020. Furthermore, our multivariable model also accounts for country-level international travel restrictions adopted in response to the spread of COVID-19.

The main limitation of the study stems from the ecological study design. Despite the fact that we did not find any association between comorbidities such as diabetes and cancer and the mean death rates at country level, it is possible for an individual with any or all of these comorbid conditions to be more susceptible to the infection and consequently at increased risk of dying. Only including countries that had reported at least 25 deaths reduced our sample and consequently the power. Furthermore, the reasonably large number of country level predictors relative to the number of countries means that we cannot rule out the potential for overfitting in the multivariable model. This may lead to spurious associations between predictors and the outcome. Other explanatory variables associated with COVID-19 related mortality may have been missed and some of the covariate data used in our model predate the COVID outbreak and may not be relevant at this time point. Furthermore, as new countries are affected by the epidemic, the virulence of the virus and resistance of the human body may have changed over time which was not accounted for in our model.I It is also possible that the quality of data, especially underreporting of deaths related to between-country differences in defining COVID-19 deaths, may have been associated with some of the predictors in our model as well as our chosen outcome and thus biased our results. Furthermore, the delay between COVID-19 symptom onset and hospitalisation may be an important factor in the overall clinical prognosis of patients with severe COVID-19 disease. Nevertheless, given that our analyses rely on country-level determinants and in the absence of individual patient data, it is impossible to ascertain the country-level trends of delay to hospital admission. Notwithstanding, some other country-level parameters pertaining to the accessibility of healthcare included in our analyses such as the number of hospital beds per 10,000 population, proportion of population living in urban areas as well as the WHO health index may account for such differences.

## CONCLUSION

Out of all the country-level parameters assessed, international travel was the main predictor of the severity of the first global wave of the COVID-19 pandemic. Given that many of world middle and lower-income countries are showing signs of continued rise in infection rates, international travel restrictions applied very early in the pandemic course should be considered to avoid rapidly increasing infection and death rates globally. The associations between other predictors, such as BCG vaccination coverage, prevalent hypertension and COVID-19 testing capacity, and the outcome were weaker and need to be interpreted in the light of our ecological study design. Further studies are required to determine the relationship between previous BCG vaccination and COVID-19 disease progression.

## Data Availability

The data utilised in this study have been uploaded as a supplementary file

## CONTRIBUTORSHIP

PKM and SB conceived the idea. TAP, DTG, ZP, WAS, JAP and KDE collected data and performed literature search. TAP, PKM, DJM and SB developed analysis plan. TAP analysed the data under supervision of DJM. TAP and SB drafted the paper. All authors contributed to the interpretation of results and in making an important intellectual contribution to the manuscript. All authors read and approved the final manuscript.

## ACKNOWLEDGEMENTS

We would like to thank Dr Kathryn Martin who provided valuable advice in study design.

## DATA SHARING STATEMENT

All data relevant to the study have been submitted to the journal as supplementary materials.

